# Data Driven Stochastic Model for Detecting Patients with Alzheimer’s Disease

**DOI:** 10.64898/2026.06.06.26355081

**Authors:** Dilmi Abeywardana, Chris P. Tsokos

## Abstract

Alzheimer’s disease (AD) is a critical neurological disorder that causes the brain to shrink and leads to the eventual death of brain cells, adversely affecting a person’s ability to function. AD is a fast-growing disease in the United States and was the fifth leading cause of death among Americans 65 years of age or older in 2023. In the United States, 6.9 million people aged 65 or older were diagnosed with AD, along with a high rate of undiagnosed patients. Thus, the objective of our study is to develop a real data-driven predictive model to identify a patient with AD based on eight risk factors: Age, Gender, ADAS-Cog13, Entorhinal, Fusiform, Intracranial Volume (ICV), Amyloid-Beta, and Tau Protein, with a high degree of accuracy. The quality of the model was evaluated using well-established and sophisticated statistical measures: the area under the receiver operating characteristic curve, calibration plot, Hosmer–Lemeshow goodness-of-fit test, and K-fold cross-validation. If a patient is given information on the above risk factors, our proposed binary logistic regression model can classify the patient as having AD or not with at least 98% accuracy.

## 1 Introduction

Alzheimer’s disease (AD) is a multifactorial neurodegenerative disease (brain disorder) that slowly destroys the memory, problem-solving, and thinking ability of a person over time. AD starts years before symptoms appear, eventually leading to loss of brain cells and connections between the brain and daily activities, and results in irreversible complex changes in the brain. In 1906, AD was first introduced by Dr. Alois Alzheimer. He noted some changes in brain tissue from a woman who died with an unrecognized mental condition, including symptoms such as memory loss, language problems, and unexpected behaviors.

Alzheimer’s disease is a fast-growing disease in the United States, and one in three Americans who are older dies due to AD or other dementia. In 2025, the estimated number of AD patients in the United States is approximately 7.2 million at the age of 65 or older, or 1 in 9 people 65 or older has AD. Among them, 74% are 75 years or older. By 2050, the estimated growth of AD patients 65 years of age or older may reach 13 million if there is no medical development to prevent or cure AD. In addition, the rate of undetected patients with AD is too high due to the lack of knowledge and the difficulty of identifying patients with AD. AD is the most common type of dementia, and about 55 million people in the world live with dementia, and 70% of dementia cases are accounted for.

The causes of AD are not fully identified and medical professionals and researchers believe that AD is caused by a combination of genetic, lifestyle, and environmental factors that affect the brain over a long period of time. They believe that a key part of AD is due to two substances within the brain called amyloid-beta and tau proteins. Amyloid-beta is a fragment of a large protein that clumps together to form plaques between neurons and disrupts communication between brain cells in a healthy person. Tau proteins support the transport of nutrients and other essential materials; however, AD causes tau proteins to change shape and structure into neurofibrillary tangles.

The main symptom of a patient with AD is memory loss, such as difficulty remembering conversations and clearly thinking, which worsens over time. They repeat statements again and again, misplace items, get lost in familiar places, eventually forget the names of family members and everyday objects, have difficulty expressing their own thoughts, and so on. In addition, AD can affect the person’s behaviors, such as depression, social withdrawal, distrust in others, loss of interest in activities, different sleeping habits, wandering, etc. However, symptoms of AD, such as changes in memory, thinking, remembering, reasoning, behavior, and social skills, are known to be the main symptoms of dementia.

AD has no cure; however, it has a few treatments that prevent or slow the development of AD. Removing the amyloid-beta of a patient with AD reduces cognitive and functional decline in early Alzheimer’s, and other treatments temporarily slow down symptoms and improve the quality of life of a person [1]. Thus, it is essential to diagnose an individual with AD as early as possible to delay or prevent its development during the early stage of AD.

Several risk factors have been identified as contributing to the development of AD. Aging is the best-known risk factor for AD, and it develops over time. Researchers found that one per thousand at age 65 to 74 was diagnosed as an AD patient, and thirty-two per thousand at age 75 to 84 were diagnosed as an AD patient [2]. Family history and genetics are also known to be risk factors for AD [3] [4]. Amyloid-beta and tau proteins also cause AD [5]. Two-thirds of females tend to have AD compared to males, making sex a contributing risk factor [6] [7]. A good education level, along with social skills, also positively affects AD. A healthy lifestyle, such as regular exercise and the management of blood pressure, diabetes, cholesterol, etc, will lead to a low risk of having AD [8] [9] [10] [11]. Thus, identifying risk factors and their contribution can lead to success in preventing AD.

Since there is no exact known cause of AD, medical professionals and researchers rely more on descriptive methods to determine whether a patient has AD or not. In the literature, we have found that univariate and multivariate logistic regression have been utilized to discriminate clinically significant Prostate cancer and for the early detection of Parkinson’s patients based on heterogeneous risk factors. These methodologies have been extensively used in medical research to improve the detection of clinically significant cases [12], [13]. Thus, the objective of this study is to develop a real data-driven binary logistic regression model to classify whether a person has AD or not, relating several risk factors: Age, Gender, ADAS-Cog13, Entorhinal, Fusiform, ICV, Amyloid-beta, and Tau proteins, with a high degree of accuracy. Furthermore, our proposed real data-driven stochastic model has identified significant individual risk factors as well as their two-way interactions that contribute significantly to AD. To our knowledge, there is no such stochastic regression model considering the contribution of interactions. In addition, the proposed model was statistically evaluated for its high degree of quality.

The chapter is organized as follows. Section 2 is about the real data that have been used to develop the model, preliminary data analysis, and the step-by-step process of model development and evaluation methods. Section 3 discusses the results of the model development and its evaluation. Finally, Section 4 concludes with the important achievements of our final analytical model by summarizing all significant findings.

## 2 Methodology

### 2.1 The Real Data

Data used in the preparation of this article were obtained from the Alzheimer’s Disease Neuroimaging Initiative (ADNI) database (adni.loni.usc.edu). The ADNI was launched in 2003 as a public-private partnership led by Principal Investigator Michael W. Weiner, MD. The primary goal of ADNI has been to test whether serial magnetic resonance imaging (MRI), positron emission tomography (PET), other biological markers, and clinical and neuropsychological assessment can be combined to measure the progression of mild cognitive impairment (MCI) and early Alzheimer’s disease (AD). The final data set consists of 311 individuals with and without Alzheimer’s disease (AD). We initially started to develop our model with fourteen risk factors and identified only eight risk factors that significantly contribute to the classification of an individual with AD or not. The figure 1, given the following, is the network of data, including categorical and continuous risk factors that we have applied for our model development process.

**Fig 1.**
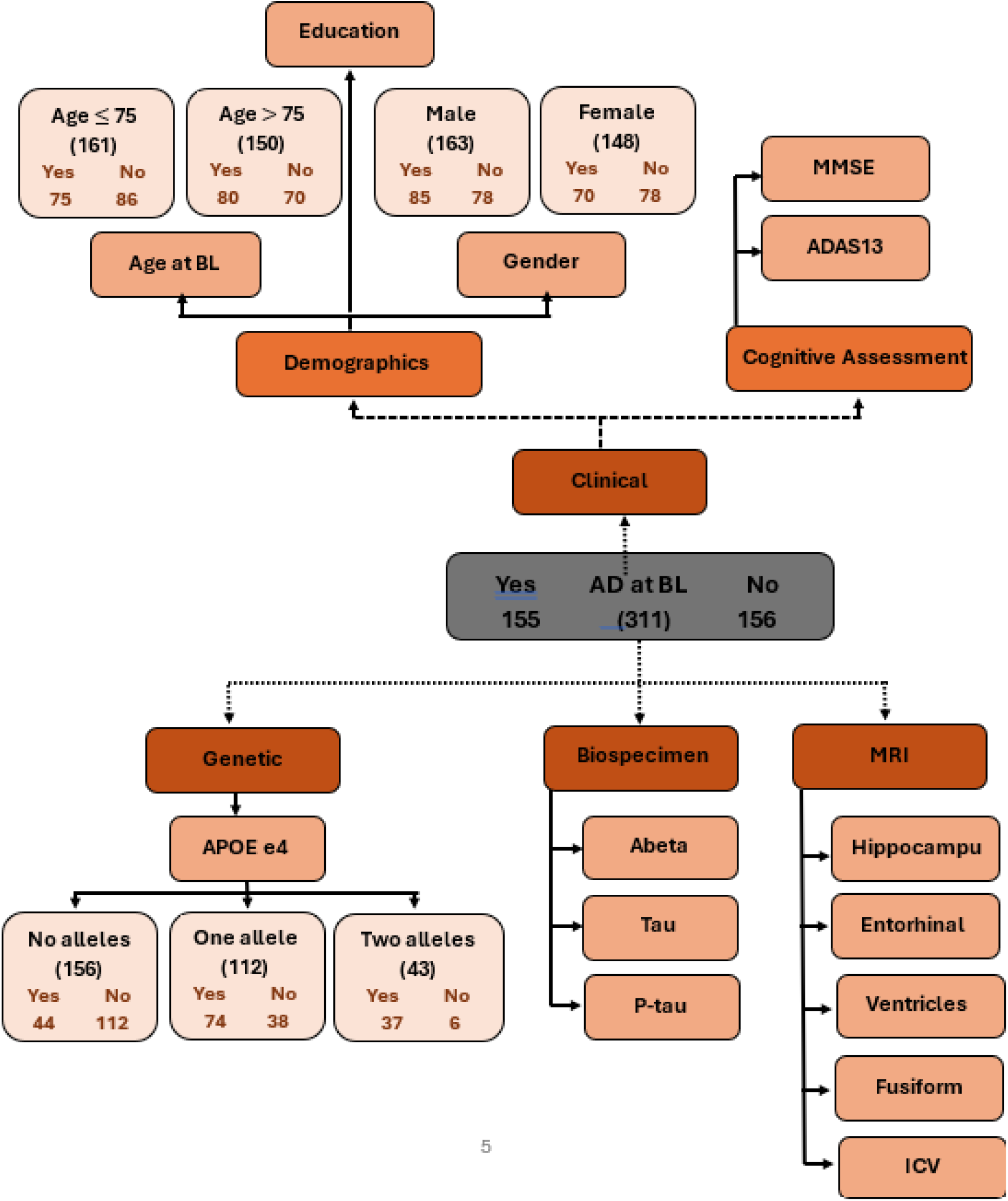
Data Diagram of Risk Factors for Individuals Having AD or Not.

The risk factor Age ranges from 55 to 90 and was separated into two categories: Age ≤ 75 and Age *>* 75. Gender has two categories: Male and Female. Apolipoprotein-E (APOE) is a categorical genotype risk factor with three categories (Non-carrier, Homozygous carrier, and Heterozygous carrier). APOE gene consists of three main alleles: *ϵ*2 (APOE2), *ϵ*3 (APOE3), and *ϵ*4 (APOE4). However, APOE *ϵ*4 is the most common risk factor for having AD by increasing the risk of developing the disease. In addition, clinical trials conduct different kinds of cognitive assessments to assess AD. We have selected two such scales: Alzheimer’s Disease Assessment Scale-Cognitive Subscale (ADAS-Cog) and Mini-Mental State Examination (MMSE) for our study. However, the ADAS-Cog is more sensitive and reliable than the MMSE because it is less influenced by educational level and language skills. Moreover, Magnetic Resonance Imaging (MRI) data are used to detect brain changes, which include volumetric measurements (*mm*^3^) of the hippocampus, Entorhinal, Ventricles, Fusiform, and Intracranial volume (IVC). Amyloid-Beta is a peptide that forms plaques around brain cells, while Tau protein forms tangles within brain cells. Amyloid-Beta, P-Tau, and Tau proteins were measured in picograms per milliliter (*pg/ml*).

Before starting the model development procedure, it is essential to identify whether there is a significant difference between the levels of categorical variables with respect to the response. Thus, we have used the Wilcoxon rank sum test with continuity correction to check the hypothesis whether there is any significant difference between the individuals being classified as having AD or not with respect to their Gender: Male and Female, and Age: ≤ 75 and *>* 75, separately. For both cases, the test failed to reject the null hypothesis (There is no significant difference with respect to gender or age in being classified as having AD or not), having p-values of 0.3941 (*>* 0.05) and 0.2353 (*>* 0.05), respectively. Thus, we continued our model development procedure without separating the gender into male and female individuals, as well as separating age groups.

### 2.2 Preliminary Statistical Analysis

Table 1, given below, provides the descriptive analysis of all continuous risk factors, including their mean, standard deviation, median, minimum, maximum, skewness, and kurtosis. These measures provide information about how our risk factors vary within a specific limit with respect to the characteristics of the risk factor. Figure 2 illustrates the correlation between individual risk factors. We can notice that there are high correlations (*>* 0.70) between individual risk factors such as **Tau protein** and **P-tau protein, ADAS-Cog13** and **MMSE** scores, and **Hippocampus** and **Entorhinal**.

**Table 1.** Descriptive Analysis for Continuous Risk Factors.

| Indicators | Mean | Std.Dev | Minimum | Maximum | Skewness | Kurtosis |
| --- | --- | --- | --- | --- | --- | --- |
| Education | 15.76 | 2.66 | 6 | 20 | -0.31 | -0.24 |
| ADAS-Cog13 | 20.06 | 12.67 | 0 | 51 | 0.46 | -0.81 |
| MMSE | 26.20 | 3.33 | 19 | 30 | -0.36 | -1.24 |
| Hippocampus | 6646.27 | 1204.56 | 3617 | 9572 | -0.15 | -0.70 |
| Entorhinal | 3324.37 | 800.73 | 1438 | 5598 | -0.06 | -0.50 |
| Ventricles | 41440.63 | 20899.49 | 7753 | 116677 | 0.96 | 0.90 |
| Fusiform | 16741.66 | 2668.73 | 10012 | 24788 | -0.04 | -0.01 |
| ICV | 1522341.67 | 175487.66 | 1100690 | 2057400 | 0.28 | -0.48 |
| Abeta | 819.97 | 367.97 | 203 | 1678 | 0.70 | -0.50 |
| P-tau | 29.26 | 15.04 | 9.56 | 83.3 | 1.13 | 1.10 |
| Tau | 299.29 | 139.19 | 107.3 | 793.4 | 1.16 | 1.29 |

**Fig 2.**
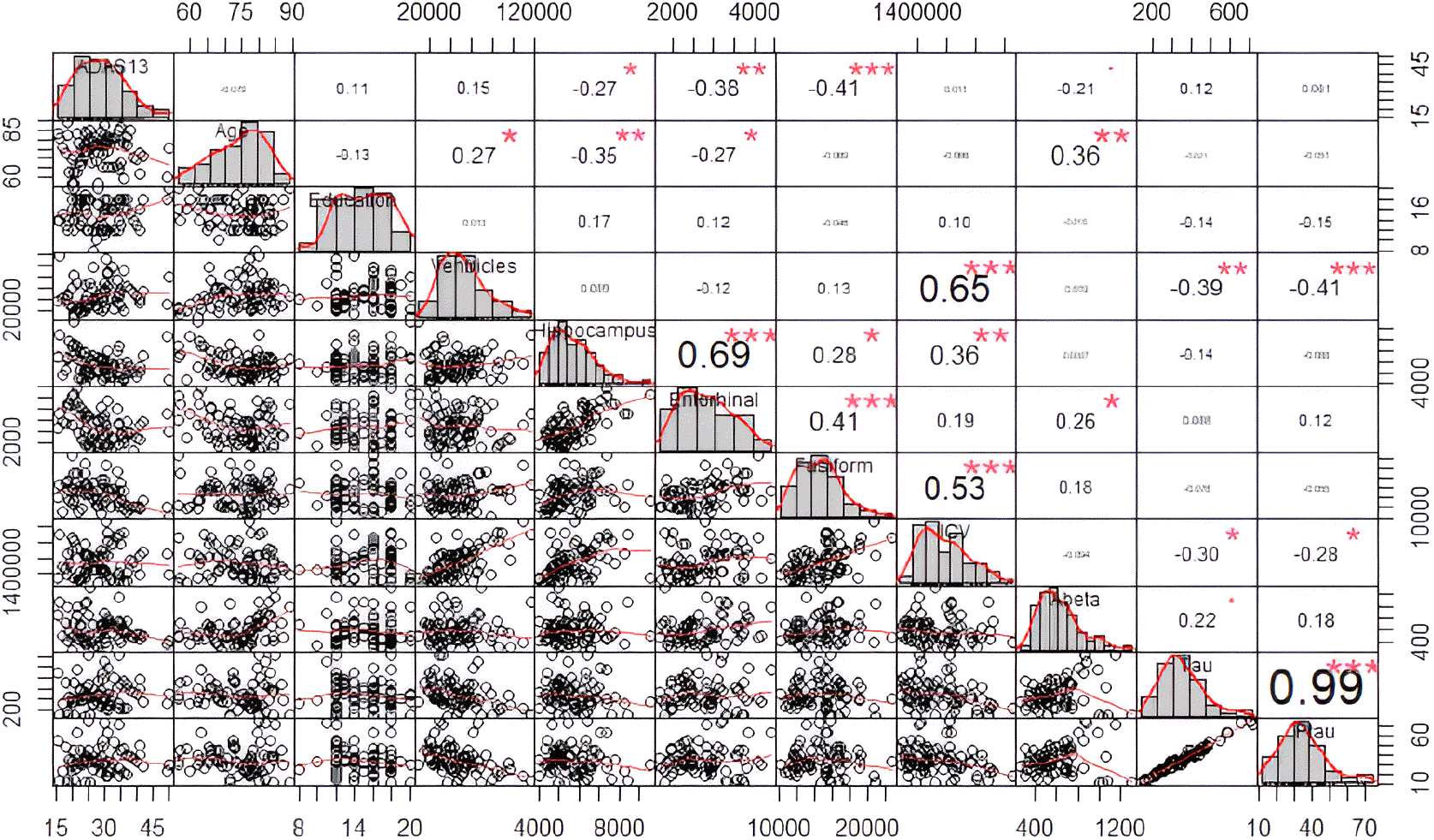
Correlation Matrix for Continuous Risk Factors.

In Table 1, above, we can clearly identify that some risk factors indicate higher variances while others have small variances. These high variations in variance may lead our developed model to conclude meaningless results due to different scales. Thus, before proceeding with model development, we addressed this issue by scaling (standardizing) all continuous risk factors to the same.

Standardization is also known as Z-score normalization, which is a commonly used technique to rescale the variables in a data set to bring them into the same scale through a mean value of zero and a standard deviation value of one, following the properties of the Gaussian probability distribution. The formula 1 given below was applied to all continuous risk factors to standardize the data.

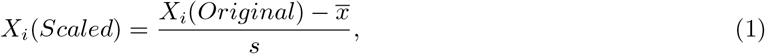

where *X_i_*(*Scaled*) is the *i^th^* standardized value, *X_i_*(*Original*) is the *i^th^* original observation value, 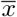 is the sample mean, and *s* is the sample standard deviation for the observations of a particular risk factor.

### 2.3 Development of the Binary Logistic Regression Model

In this study, our main goal is to develop a real data-driven binary logistic regression model that classifies whether an individual has AD or not, considering the statistically significant contributing individual risk factors and their two-way interactions with a high degree of accuracy.

A logistic regression model, also known as the logit model, is used to model the log-odds of an event as a linear combination of one or more independent variables. In the binary logistic regression model, the response is dichotomous, that is, the outcome is limited to two possible outcomes, either 0 or 1, while the independent variables are continuous or categorical.

The general analytical form of the logistic regression model with all the individual risk factors and two-way interactions can be expressed as follows,

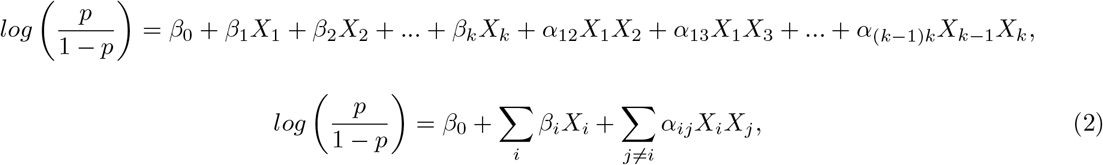

where *p* is the probability of success, *β*_0_ is the intercept of the linear regression model, *X_i_* is the *i^th^* individual risk factor, *β_i_* is the weight of the *i^th^* individual risk factor, *α_ij_* is the weight of the interaction between *i^th^* and *j^th^* individual risk factors.

This generalized regression model (equation 2) predicts the values for the logit function, that is, the natural log of the odds. Thus, we need to convert the output using equation 3, given below, to get the corresponding probability value of the response.

The statistical form of the binary logistic function can be expressed as follows,

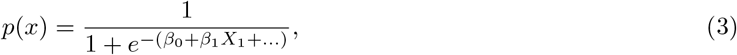

where *e* is the base natural logarithm and *p*(*x*) is the probability [0, 1] of the response variable (success). If the probability is less than a predefined threshold (0.5), then we predict the response as 0, and if the probability is greater than or equal to that predefined threshold (0.5), we predict the response as 1.

There are three key properties of a logistic regression. They are,

- The response variable should obey the Bernoulli distribution (only two outcomes).
- Parameter estimates are based on maximum likelihood estimation.
- The coefficient of determination (*R*^2^) is not a suitable measure to evaluate the model performance.

### 2.4 Validation of the Predictive Model

An analytical model has been developed on one or more assumptions, and it is essential to satisfy the underlying assumptions of the corresponding model to obtain accurate results. The following are the key assumptions that must be satisfied by a logistic regression model.

1. The response variable should be binary or dichotomous.
2. There should be little or no multicollinearity among risk factors (i.e., Risk factors should be independent of each other). This can be addressed by using the Variance Inflation Factor (VIF) values (VIF *<* 10).
3. There should be linear relationships between the risk factors and the log odds.
4. There should not be extreme outliers or influential points.

More details on this subject area can be found in the literature [14] [15] [16].

### 2.5 Evaluation of the Predictive Model

We have utilized the methods: Hosmer and Lemeshow test, confusion matrix, sensitivity analysis, ROC curve and AUC score, calibration curve, K-fold cross-validation, information criteria such as AIC, and residual plots to evaluate the high quality and performance of our proposed binary logistic regression model.

#### 1. Hosmer and Lemeshow Test (HL Test)

The Hosmer and Lemeshow Test is a statistical test that assesses the goodness of fit of a logistic regression model and only works if the response is binary [16].

First, we regroup (g number of groups) the observations by ordering the predicted probabilities and calculate the HL test statistic using the following equation,

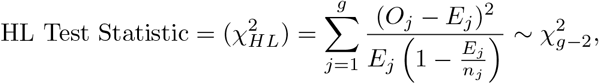

where, g is the number of groups, *O_j_* is the observed frequency of the *j^th^* group, *E_j_* is the expected frequency of the *j^th^* group and *n_j_* is the number of observations in the *j^th^* group.

The test results have two outcomes: the HL chi-squared value and the p-value. If the p-value is too small (*<*0.05), then the fit of the model is not a better fit.

#### 2. Confusion Matrix

The confusion matrix is a contingency table that includes details on the performance of a classification model. It provides the accuracy of the model’s predictions by comparing them with actual values.

**Table 2.** Contingency Table for Confusion Matrix.

| Actual | Predicted |  |
| --- | --- | --- |
|  | Positive | Negative |
| Positive<br>P | True Positive<br>TP | False Negative<br>FN |
| Negative<br>N | False Positive<br>FP | True Negative<br>TN |

where TP means that the model accurately predicts the positive class, FP means that the model incorrectly predicts the positive class, TN means that the model accurately predicts the Negative class, and FN means that the model incorrectly predicts the negative class.

This confusion matrix can be utilized to derive three concepts: **Sensitivity, Specificity**, and **Accuracy** of a logistic model to evaluate its performance [17] [18].

##### Sensitivity

Sensitivity is the true positive rate, that is, the proportion of the number of positive cases that the model correctly predicted.

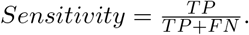

##### Specificity

Specificity is the true negative rate, that is, the proportion of the number of negative cases that the model correctly predicted.

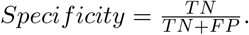

##### Accuracy

Accuracy is the percentage of correct predictions.

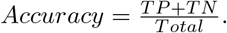

#### 3. Receiver Operating Characteristic (ROC) Curve and Area Under Curve (AUC)

The Receiver Operating Characteristic (ROC) curve is a graphical visualization of a binary classifier between sensitivity and specificity. It plots the true positive rate (sensitivity) against the false positive rate (1-specificity) at various probability thresholds. The Area Under Curve (AUC) value is the area between the ROC curve and the x-axis. This measure can be used to evaluate the performance of a binary logistic model, where the quality of the model is better with a higher AUC value.

#### 4. Calibration Curve

The calibration curve is a graphical visualization to present the connection between the estimated and observed probabilities. If the model is well calibrated, then the predicted probability should be close to the real probability of the event. Thus, a good fit indicates that the points of the calibration curve follow the diagonal line.

#### 5. K-fold Cross-Validation

The k-fold cross-validation is a commonly used statistical model validation method that evaluates the performance of a statistical model. The k-fold cross-validation method randomly divides the data into k subsets and treats k-1 subsets as the training sets while keeping the remaining one as the test set. Then the model is fitted using k-1 subsets (training set) and validates its performance using the test set. Since we have k subsets, we repeat this procedure k times until every sample serves as a test set, and take the average performance for all k validations. Figure 3 provides an overview of the process of the k-fold cross-validation method. More information can be found in the literature [19], [20] [21].

**Fig 3.**
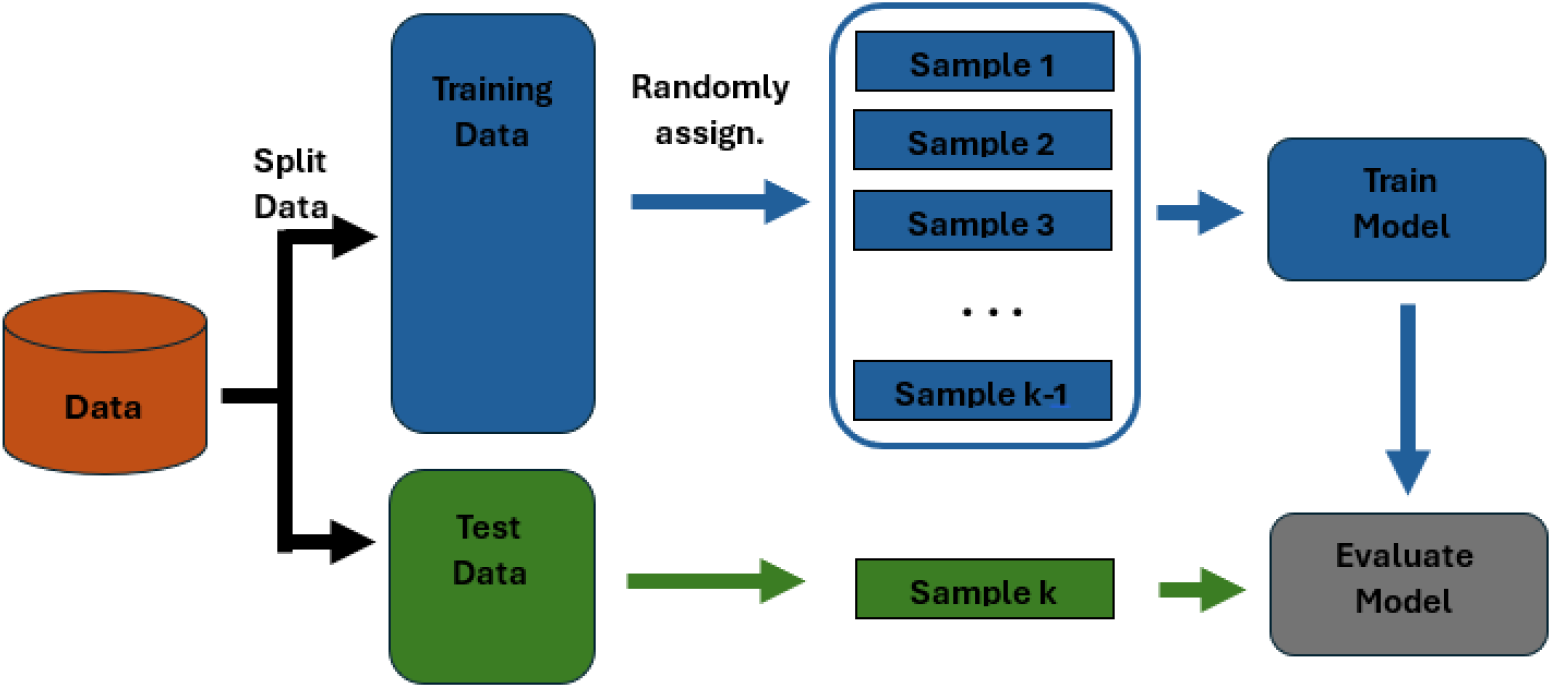
Overview of K-Fold Cross Validation.

## 3 Important Results

In Section 2.2, we have identified that the variances of some risk factors are too high and some are too small. Thus, we need to bring them to the same scale before developing the logistic regression model. We have applied the standardization method to our continuous risk factors, and Figure 4, given below, presents the corresponding parameters (mean and standard deviation) for each continuous risk factor used during the standardization process.

**Fig 4.**
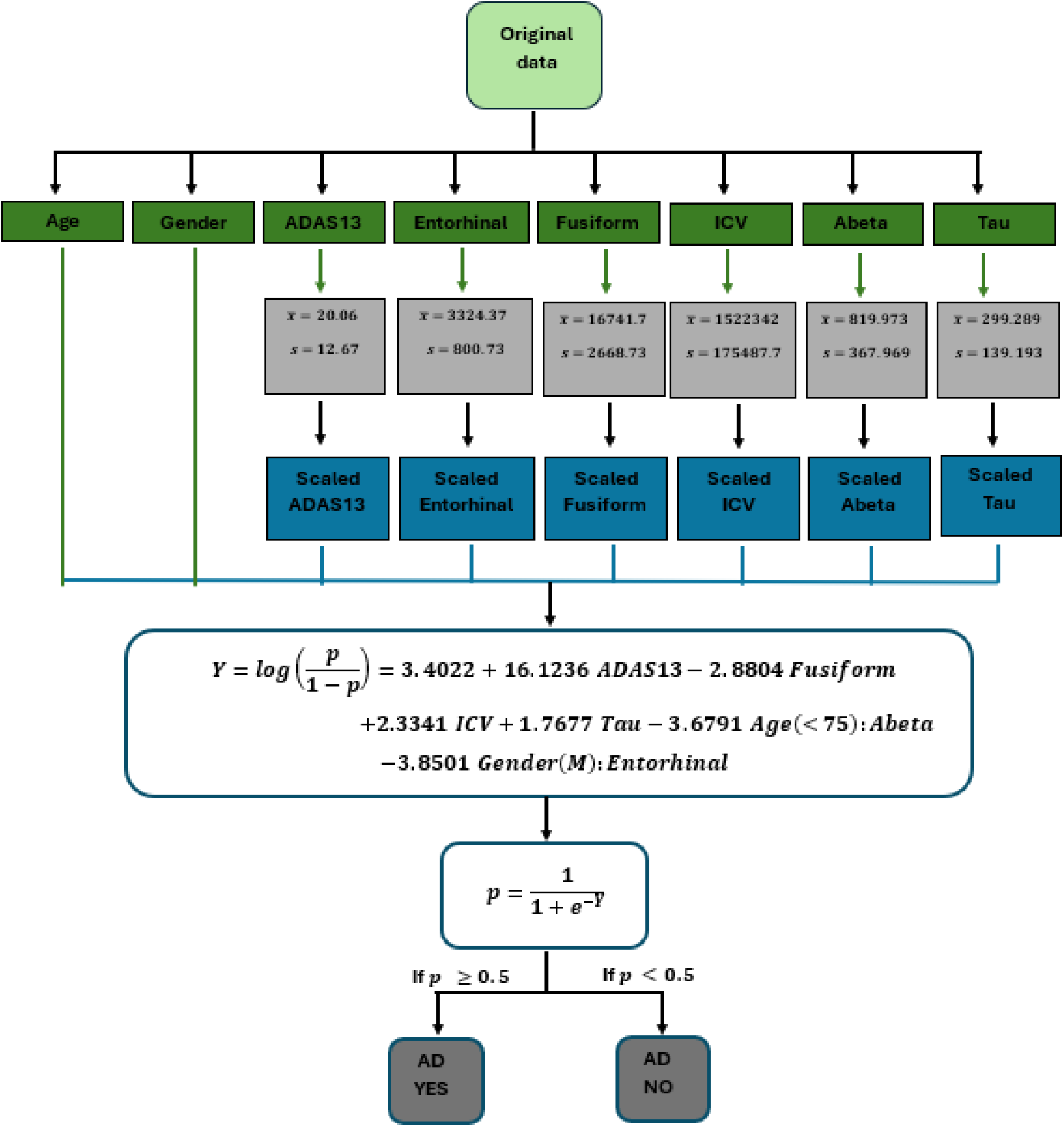
Architecture for Classifying a Patient with AD or Not.

Then we divided our data into training (80%) and test (20%) sets with 249 and 62 observations, respectively. The training data set was used to develop the binary logistic regression model discussed in Section 2.3. The backward stepwise elimination method with a stopping rule based on Akaike’s information criterion (AIC) was conducted to select the significant risk factors and their two-way interactions. The analytical form of the final model consists of four individual risk factors and two interacting terms that significantly contribute to the response as given below,

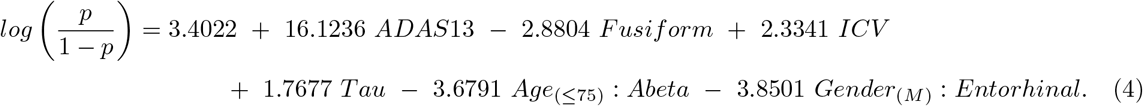

Suppose that an individual can provide the necessary information on risk factors, given in Figure 4. In that case, our proposed stochastic model can predict whether that individual has AD or not with at least **98%** degree of accuracy. According to the confusion matrix (Table 3), given below, the **sensitivity** and the **specificity** of our proposed model are **96.80%** and **98.37%**, respectively, validating the high quality of our proposed model.

**Table 3.** Confusion Matrix for Training Set.

| Actual | Predicted |  |
| --- | --- | --- |
|  | Positive | Negative |
| Positive | True Positive<br>121 | False Negative<br>4 |
| Negative | False Positive<br>2 | True Negative<br>121 |

The following Figure 5 graphically illustrates the predicted probabilities of being AD or not for the individuals in our training set.

**Fig 5.**
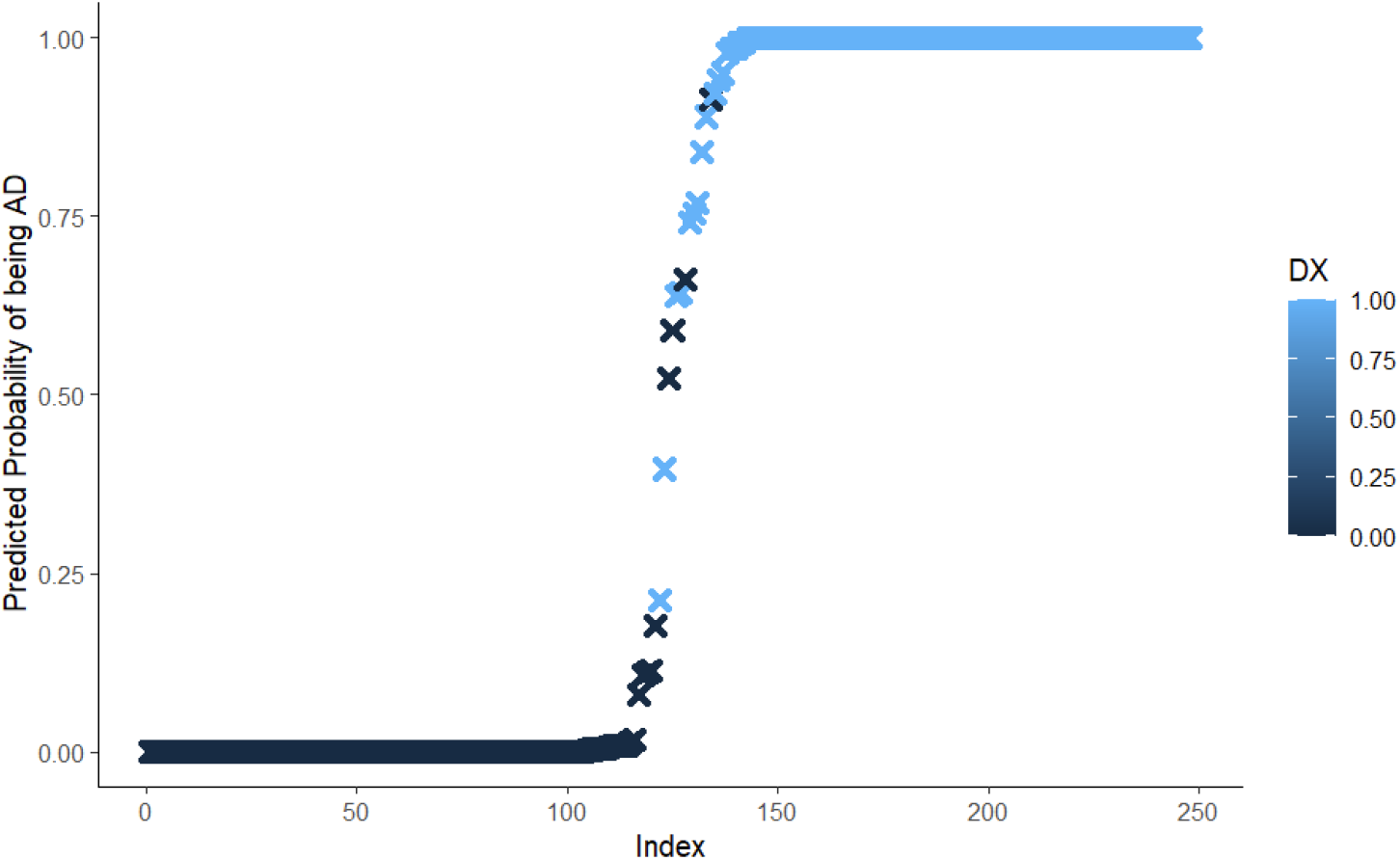
Predicted Probabilities of Being AD.

The Akaike information criterion (AIC) value for the proposed predictive model is 35.309, a small value that confirms the high quality of our model.

The Hosmer-Lemeshow test value was calculated to check the goodness of fit of the proposed predictive model, and the p-value is 0.99 (*>*0.05), confirming that our proposed model is a perfect fit for the training data at a 95% level of significance.

In addition, we graphically visualized the calibration curve, given in Figure 6, for our proposed model, and it follows the diagonal line. This indicates that the proposed predictive model has empirical probabilities that are close to the estimated probabilities, and the calibration is well-calibrated with a good fit.

**Fig 6.**
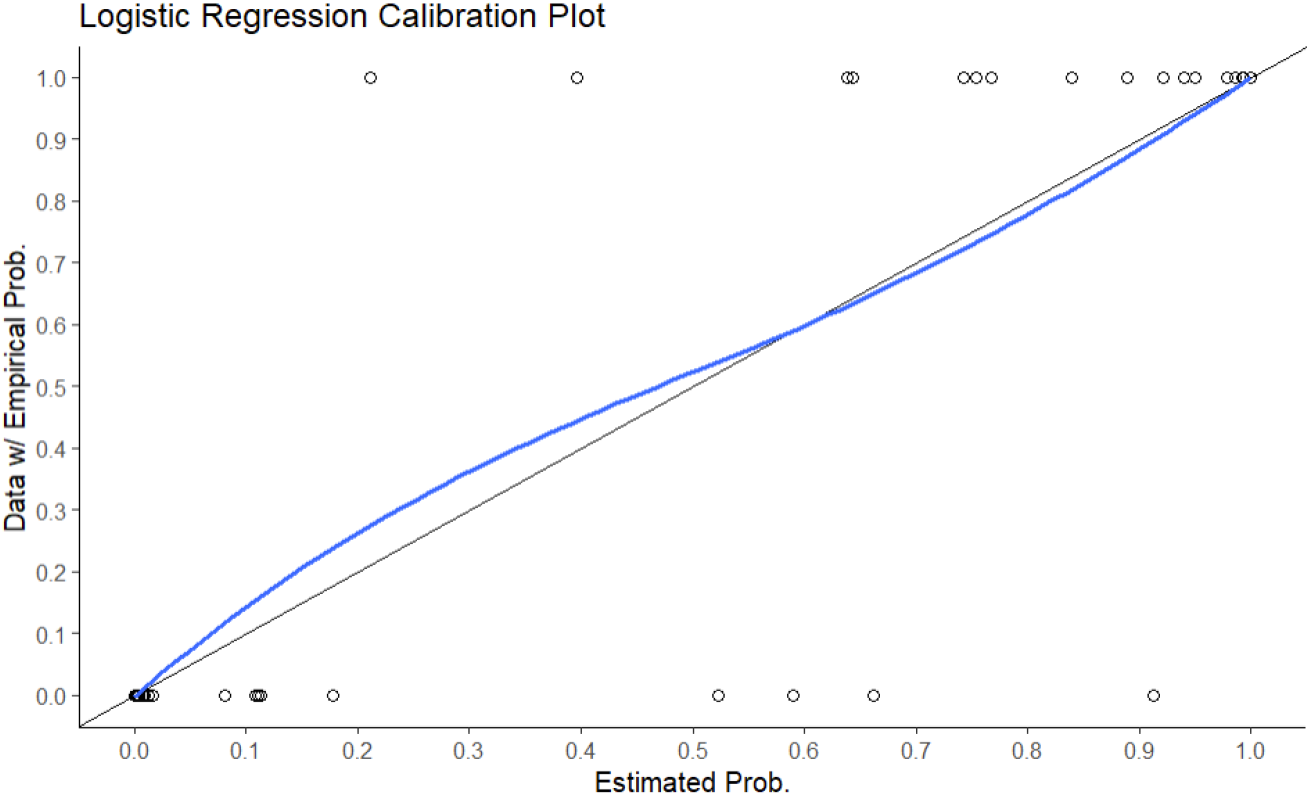
Calibration Curve.

Figure 7 provides the graphical interpretation of the receiver operating characteristic (ROC) curve and the area under the curve (AUC) value for the proposed predictive model. The AUC value is 0.999, a very high value confirming the high quality of our proposed predictive model.

**Fig 7.**
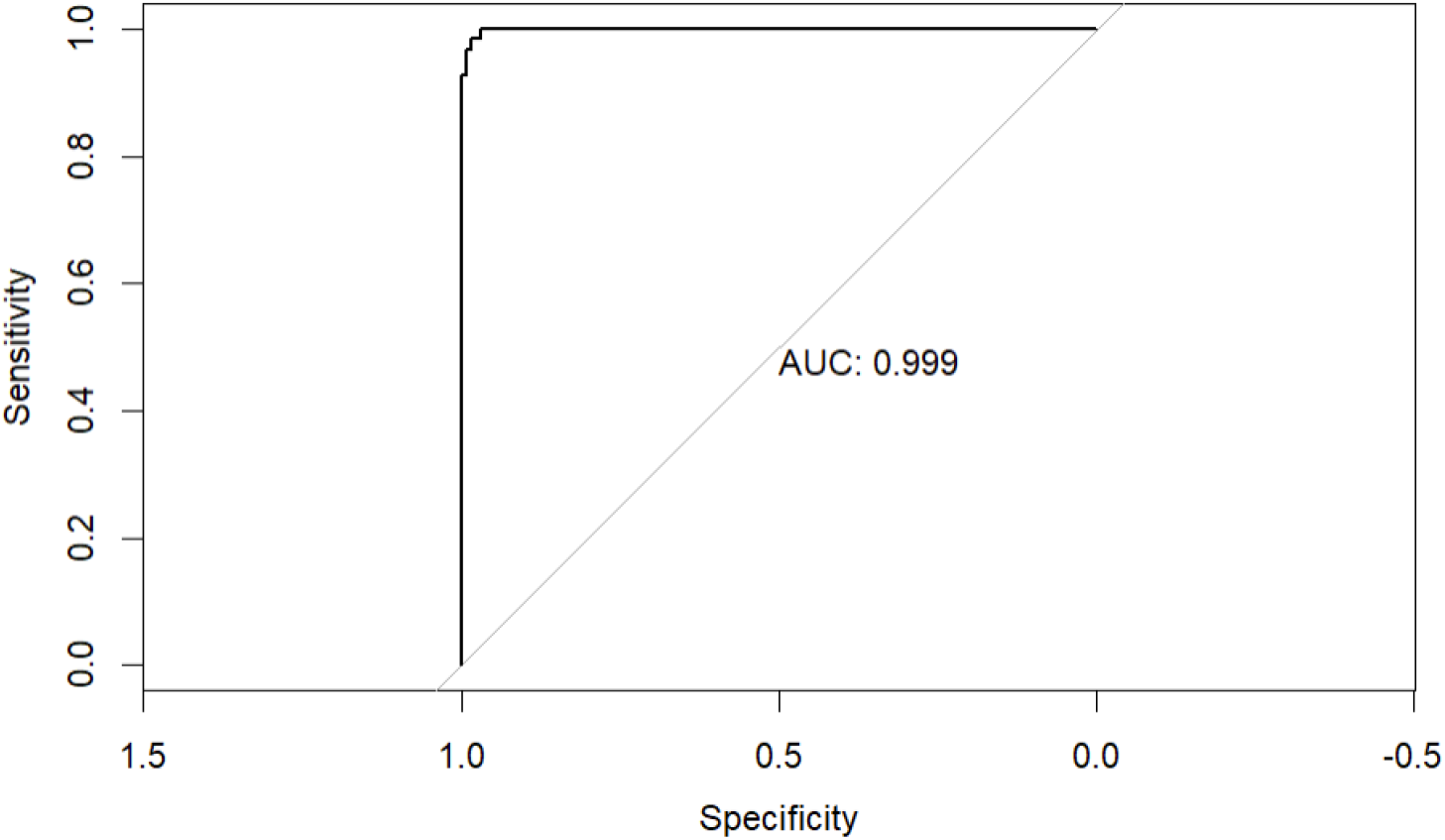
ROC Curve with AUC Value.

Moreover, we checked the assumptions on the proposed binary logistic regression model one by one, and our model has satisfied all the assumptions discussed in Section 2.4.

In our proposed predictive model, there is no multicollinearity among the risk factors. Table 4, below, provides the VIF values for each risk factor, and all values are small, less than 10, confirming the fact that there is no multicollinearity among risk factors.

**Table 4.**
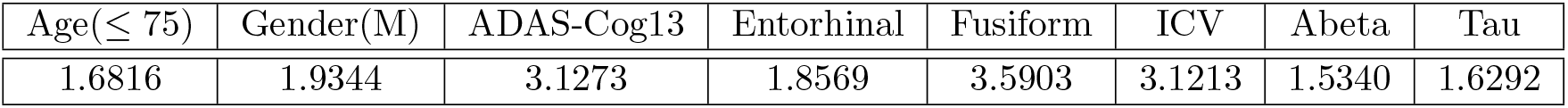
VIF Values for Risk Factors.

| Age($\leq 75$) | Gender(M) | ADAS-Cog13 | Entorhinal | Fusiform | ICV | Abeta | Tau |
| --- | --- | --- | --- | --- | --- | --- | --- |
| 1.6816 | 1.9344 | 3.1273 | 1.8569 | 3.5903 | 3.1213 | 1.5340 | 1.6292 |

In addition, we checked for the extreme outliers and identified that there are no extreme outliers or influential points in our proposed predictive model. Figure 8 clearly illustrates the behavior of the outliers, and they are not extreme.

**Fig 8.**
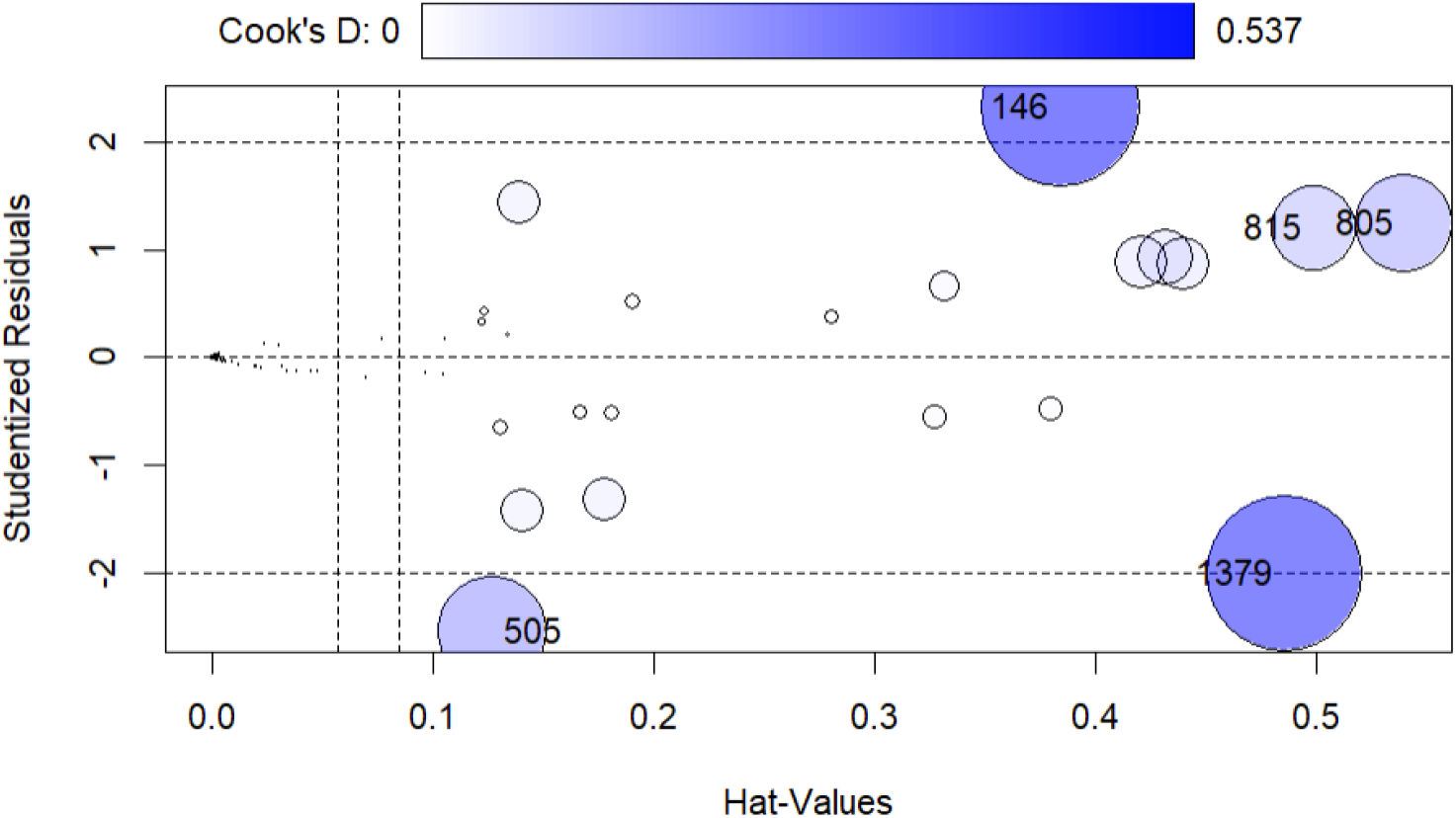
Cook’s D Values for Checking Outliers.

The graphical visualization in Figure 9 clearly illustrates the linear relationship between each continuous risk factor: Abeta, Tau-Protein, ADAS13, Entorhinal, Fusiform, ICV, and the log odds.

**Fig 9.**
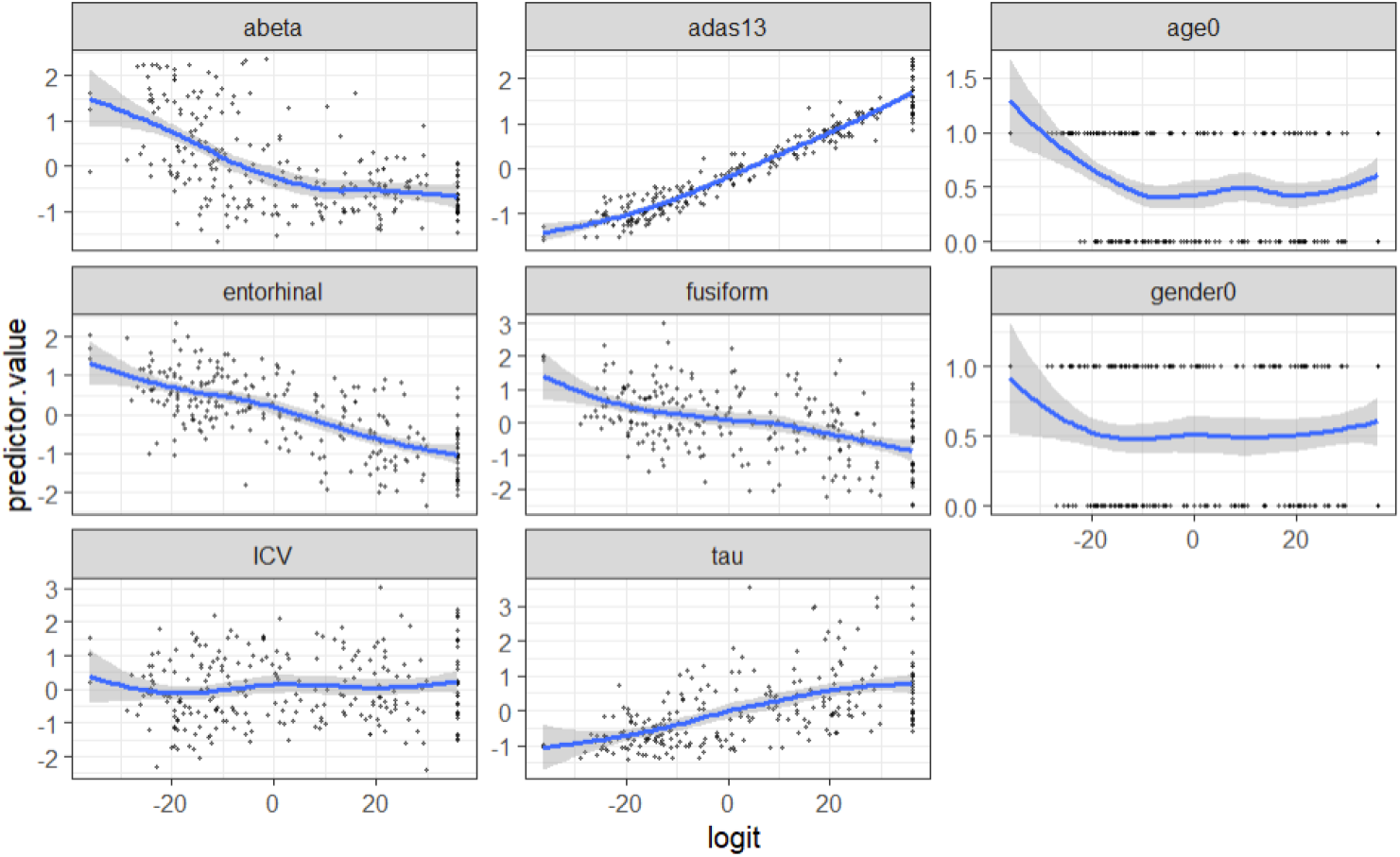
Linearity Between Continuous Risk Factors and Log Odds.

Since our proposed model satisfied all the assumptions made on binary logistic regression, we applied the K-fold cross-validation (10-fold) technique to validate the model performance as given in Table 5, below.

**Table 5.** Evaluation Results of 10-Fold Cross Validation.

| Training set |  |  |  | Test set |  |  |  |
| --- | --- | --- | --- | --- | --- | --- | --- |
| Accuracy | Kappa | Sensitivity | Specificity | Accuracy | Kappa | Sensitivity | Specificity |
| 97.58% | 0.9516 | 96.80% | 98.37% | 98.39% | 0.9677 | 100% | 96.77% |

The accuracy represents the percentage of total correctly classified cases out of all classifications. Kappa is a statistic for accuracy, which accounts for baseline probabilities from the null model (without predictors).

## 4 Usefulness and Contribution

In this study, we have developed a real data-driven stochastic model to predict if an individual has **Alzheimer’s disease** or not for a given set of risk factors with a high degree of **98%** accuracy.

We have identified four individual risk factors that statistically significantly contribute to the cause of having AD or not. They are,

1. ADAS-Cog13
2. Fusiform
3. ICV
4. Tau Protein

We have identified two interacting terms of the individual risk factors that statistically significantly contribute to the cause of having AD or not. They are,

1. Age(≤ 75) : Abeta
2. Gender(M) : Entorhinal

According to the proposed predictive model, the importance of each risk factor and interacting terms are given in Table 6, below. Using this information, medical professionals can identify the most contributing risk factor to AD and work to control its development at an early stage.

**Table 6.** Importance of the Model Terms.

| Term | Importance |
| --- | --- |
| ADAS13 | 2.884553 |
| Fusiform | 2.212239 |
| Age($\leq 75$) : Abeta | 2.082091 |
| Tau | 1.987225 |
| ICV | 1.956164 |
| Gender (M): Entorhinal | 1.915429 |

If a patient can provide information on the selected significant risk factors, then the proposed predictive model can classify whether that patient has AD or not with at least 98% accuracy. Thus, developing predictive models with respect to risk factors will be beneficial to medical professionals to identify patients with AD without depending only on descriptive methods and considering other risk factors as a combination. Moreover, the predictive model may indicate the relationship of each risk factor to the response.

Finally, all assumptions and evaluation methods, such as Confusion Matrix, ROC Curve and AUC Value, Calibration Curve, Information Criteria (AIC), and K-Fold Cross Validation, among others, have been addressed, and they uniformly confirmed the high quality of our proposed stochastic predictive model.

## Data Availability

Data used in the preparation of this article were obtained from the Alzheimer s Disease Neuroimaging Initiative (ADNI) database (adni.loni.usc.edu). The ADNI was launched in 2003 as a public-private
partnership led by Principal Investigator Michael W. Weiner, MD. The primary goal of ADNI has been to test whether serial magnetic resonance imaging (MRI) positron emission tomography (PET) other
biological markers and clinical and neuropsychological assessment can be combined to measure the progression of mild cognitive impairment (MCI) and early Alzheimer s disease (AD).

## 5 Statements and Declarations

### Ethical approval

- We, all authors, Dr. Dilmi Abeywardana and Dr. Chris P. Tsokos, have agreed on authorship, read and approved the manuscript, and given consent for submission and subsequent publication of the manuscript. Dr. Abeywardana acts as the corresponding author for this manuscript.
- We ensure that the manuscript and its contents are not a duplicate publication, and it is completely original.
- We ensure that the manuscript represents our original work and meets the ethical standards set by the Committee on Publication Ethics (COPE).

### Competing interests

The authors declare that they have no competing interests.

### Author Contributions

Conceptualization: Dilmi Abeywardana; methodology: Dilmi Abeywardana; software: Dilmi Abeywardana; validation: Dilmi Abeywardana, and Chris Tsokos; formal analysis: Dilmi Abeywardana; investigation, Dilmi Abeywardana; original draft preparation: Dilmi Abeywardana; writing review and editing: Dilmi Abeywardana and Chris Tsokos; visualization: Dilmi Abeywardana; supervision, Chris Tsokos; All authors have read and agreed to the published version of the manuscript.

### Conflicts of interest

The author(s) declared no potential conflicts of interest with respect to the research, authorship, and/or publication of this article.

## Acknowledge

Data collection and sharing for this project was funded by the Alzheimer’s Disease Neuroimaging Initiative (ADNI) (National Institutes of Health Grant U01 AG024904) and DOD ADNI (Department of Defense award number W81XWH-12-2-0012). ADNI is funded by the National Institute on Aging, the National Institute of Biomedical Imaging and Bioengineering, and through generous contributions from the following: AbbVie, Alzheimer’s Association; Alzheimer’s Drug Discovery Foundation; Araclon Biotech; BioClinica, Inc.; Biogen; Bristol-Myers Squibb Company; CereSpir, Inc.; Cogstate; Eisai Inc.; Elan Pharmaceuticals, Inc.; Eli Lilly and Company; EuroImmun; F. Hoffmann-La Roche Ltd and its affiliated company Genentech, Inc.; Fujirebio; GE Healthcare; IXICO Ltd.; Janssen Alzheimer Immunotherapy Research & Development, LLC.; Johnson & Johnson Pharmaceutical Research & Development LLC.; Lumosity; Lundbeck; Merck & Co., Inc.; Meso Scale Diagnostics, LLC.; NeuroRx Research; Neurotrack Technologies; Novartis Pharmaceuticals Corporation; Pfizer Inc.; Piramal Imaging; Servier; Takeda Pharmaceutical Company; and Transition Therapeutics. The Canadian Institutes of Health Research is providing funds to support ADNI clinical sites in Canada. Private sector contributions are facilitated by the Foundation for the National Institutes of Health (www.fnih.org). The grantee organization is the Northern California Institute for Research and Education, and the study is coordinated by the Alzheimer’s Therapeutic Research Institute at the University of Southern California. ADNI data are disseminated by the Laboratory for Neuro Imaging at the University of Southern California.

## Notes

### Competing Interest Statement

The authors have declared no competing interest.

